# Nosocomial RSV-related in-hospital mortality in children <5 years: a global case series

**DOI:** 10.1101/2022.01.10.22268872

**Authors:** Yvette N. Löwensteyn, Joukje E. Willemsen, Natalie I. Mazur, Nienke M. Scheltema, Nynke C. J. van Haastregt, Amber ten Buuren, Ichelle van Roessel, Dunja Scheepmaker, Harish Nair, Peter M. van de Ven, Louis J. Bont, RSV GOLD study group

## Abstract

**Background:** According to the World Health Organization the global burden of nosocomial infections is poorly characterized as surveillance systems for nosocomial infection are lacking. Nosocomial infections occur at higher rates in low- and lower-middle-income countries (LMICs) than in high-income countries (HICs). Current global RSV burden estimates are largely based on community-acquired disease. We aimed to characterize children with nosocomial RSV-related mortality and to understand the potential impact of RSV immunization strategies.

**Methods:** RSV GOLD is a global registry of children younger than 5 years who died with laboratory-confirmed RSV infection. We compared clinical and demographic characteristics of children with nosocomial and community-acquired RSV in-hospital mortality.

**Results:** We included 231 nosocomial and 931 community-acquired RSV-related in-hospital deaths from 65 countries. Median age at death was similar for both groups (5.4 vs 6 months). As expected, a higher proportion of children with nosocomial infection had comorbidities (87% vs 57%; p<0.001) or was born preterm (46% vs 24%; p<0.001) than children with community-acquired infection. The proportion of nosocomial deaths among all RSV deaths was lower in LMICs than in upper-middle-income countries (UMICs) and HICs (12% vs 18% and 26%, respectively).

**Conclusions:** This is the first global case series of children dying with nosocomial RSV infection. Future infant-targeted immunization strategies can prevent the majority of nosocomial RSV-related deaths. Although nosocomial RSV deaths are expected to occur at highest rates in LMICs, the number of reported nosocomial RSV deaths was low in these countries. Hospital-based surveillance is needed to capture the full burden of nosocomial RSV mortality in LMICs.

**Key points:**

- The proportion of reported nosocomial RSV-related deaths is substantially lower in lower-middle-income countries than in upper-middle-income countries and high-income countries (12% vs 18% and 26%, respectively).
- The majority of nosocomial RSV-related deaths can be prevented by infant-targeted immunization strategies as more than half were younger than 6 months of age.

## INTRODUCTION

Respiratory syncytial virus (RSV) is the leading cause of lower respiratory tract infection (LRTI) and LRTI-related death in young children. In 2015, the overall RSV mortality burden was estimated at 118200 annual deaths in children younger than 5 years of age worldwide.[1] These global burden estimates are largely based on community-acquired RSV-related deaths due to the paucity of data on healthcare-associated or nosocomial RSV-related mortality. According to the World Health Organization (WHO) the burden of nosocomial infections is poorly characterized in LMICs as surveillance systems are lacking.[2] Nevertheless, available data show that the burden of nosocomial infections is significantly higher in low- and middle-income countries (LMICs) compared to high-income countries (HICs).[3] Furthermore, RSV has been reported as the most important viral cause of nosocomial LRTI in children.[4-7]

The proportional contribution of nosocomial RSV mortality to the global RSV mortality burden is expected to be substantial, as nosocomial RSV infection is generally more severe than community-acquired RSV infection.[8-10] Currently, several RSV vaccine candidates and monoclonal antibodies are in late phase clinical development.[11] To identify vaccine target populations and to estimate the potential impact of these RSV prevention strategies, healthcare policy makers require information on the characteristics of children with life-threatening RSV infection including those with nosocomial RSV infection. However, previous reports of pediatric nosocomial RSV-related deaths are from single centres and not large enough (number of deaths ranging between 1-21)[12] to understand the demographic and clinical characteristics of children dying with nosocomial RSV infection globally.

Through our global RSV mortality registry, we aimed to describe demographic and clinical characteristics of children younger than 5 years of age dying in the hospital with laboratory-confirmed nosocomial RSV infection.

## METHODS

### Study design and patients

The RSV GOLD study is an ongoing global online mortality registry that includes individual patient data of children younger than 5 years of age who died with RSV infection. The first study results, published in 2017, included only community-acquired, in-hospital deaths.[13] Since then, the registry has been extended to also include nosocomial and out-of-hospital pediatric RSV-related deaths from January 1, 1995 onwards.[14] To obtain nosocomial RSV-related mortality patient data, we performed a systematic search in PubMed using the following search terms: “RSV” or “bronchiolitis” combined with “healthcare-associated pneumonia”, “nosocomial”, “community-acquired”, “in-hospital”, “hospital-borne”, or “cross-infection” (Supplemental Table 1). We invited authors to share patient data until July 31st, 2021 using a link to our electronic data capture platform Research Online.[15] We included published and unpublished data of children aged 0-59 months who had died with laboratory-confirmed RSV infection in the hospital between January 1, 1995 and July 31st, 2021. We excluded children for whom it was unknown where RSV infection had been acquired (in-community or in-hospital [n=489]). We collected demographic and clinical characteristics using an online questionnaire within Research Online.

### Definitions

We categorized country of origin as LMIC (LIC and LMIC combined), upper-middle income country (UMIC), or HIC according to the World Bank classifications for 2021.[16] We defined nosocomial RSV infection as a positive laboratory-confirmed RSV test in combination with a ≥48-hour timeframe between hospital admission and onset of respiratory symptoms, and as reported by the RSV GOLD collaborator. Length of hospital stay was defined as the duration of the hospital admission in days starting from the beginning of respiratory symptoms. Comorbidity was defined as the presence of at least one underlying disease. Prematurity was defined as gestational age less than 37 weeks. If data for comorbidities or prematurity were not reported (n=17), we assumed that the children were otherwise health and born at term. If data for location of death were missing (n=3), we assumed that the death had occurred in hospital.

### Nosocomial RSV infection and community-acquired RSV-related mortality

We compared the demographic and clinical characteristics of children with nosocomial RSV-related in-hospital death and those with community-acquired RSV-related in-hospital death, including previously published RSV GOLD data.[13, 17] We analyzed the data separately for different income groups and calculated for each income group the odds ratios and 95% confidence interval (CI) for the presence of demographic and clinical characteristics in children with hospital-acquired RSV relative to children with community-acquired RSV.

### Sensitivity analyses

We excluded deaths with missing data for comorbidities or prematurity and compared demographic and clinical characteristics. We also analyzed the age distribution for nosocomial RSV deaths from LMICs after excluding deaths collected from a post-mortem study conducted in Zambia in which only children aged <6 months were included (n=12).[18]

### Ethical approval

The institutional research board of the University Medical Centre Utrecht waived the requirement for parental consent as only de-identified secondary patient data are used in the RSV GOLD study. Collaborators sharing data were encouraged to adhere to local site regulations for ethics approval. Where required, local ethical approval was obtained.

### Statistical analysis

We present descriptive statistics for all variables. For continuous variables we report the median with interquartile range (IQR); for categorical variables we report the frequency and proportion. We used the Fisher’s exact test to determine statistical significance between groups for dichotomous parameters and the Mann-Whitney-U test for all continuous parameters. We applied the Bonferroni correction for multiple testing between World Bank Income groups. All statistical analyses were performed with SPSS (version 21.0; IBM Corp, Armonk, NY). The forest plot with odds ratios and 95% CIs was made in R version 4.0.3.

## RESULTS

We included a total of 231 nosocomial RSV-related deaths from 38 countries (Figures 1-2, Supplemental Table 3) of which 80 deaths were identified through the literature search (Supplemental Tables 1-2) and 188 comprised unpublished reports shared by collaborators. The 231 deaths accounted for 20% of all deaths reported to the RSV GOLD registry with available data on where RSV infection was acquired. The majority (n=123; 53%) of reported deaths were from HICs. The proportion of reported nosocomial deaths among all RSV-related in-hospital deaths was substantially lower in LMICs (12%) than in UMICs (18%, p=0.03) or HICs (26%, p<0.001).

**Figure 1:**
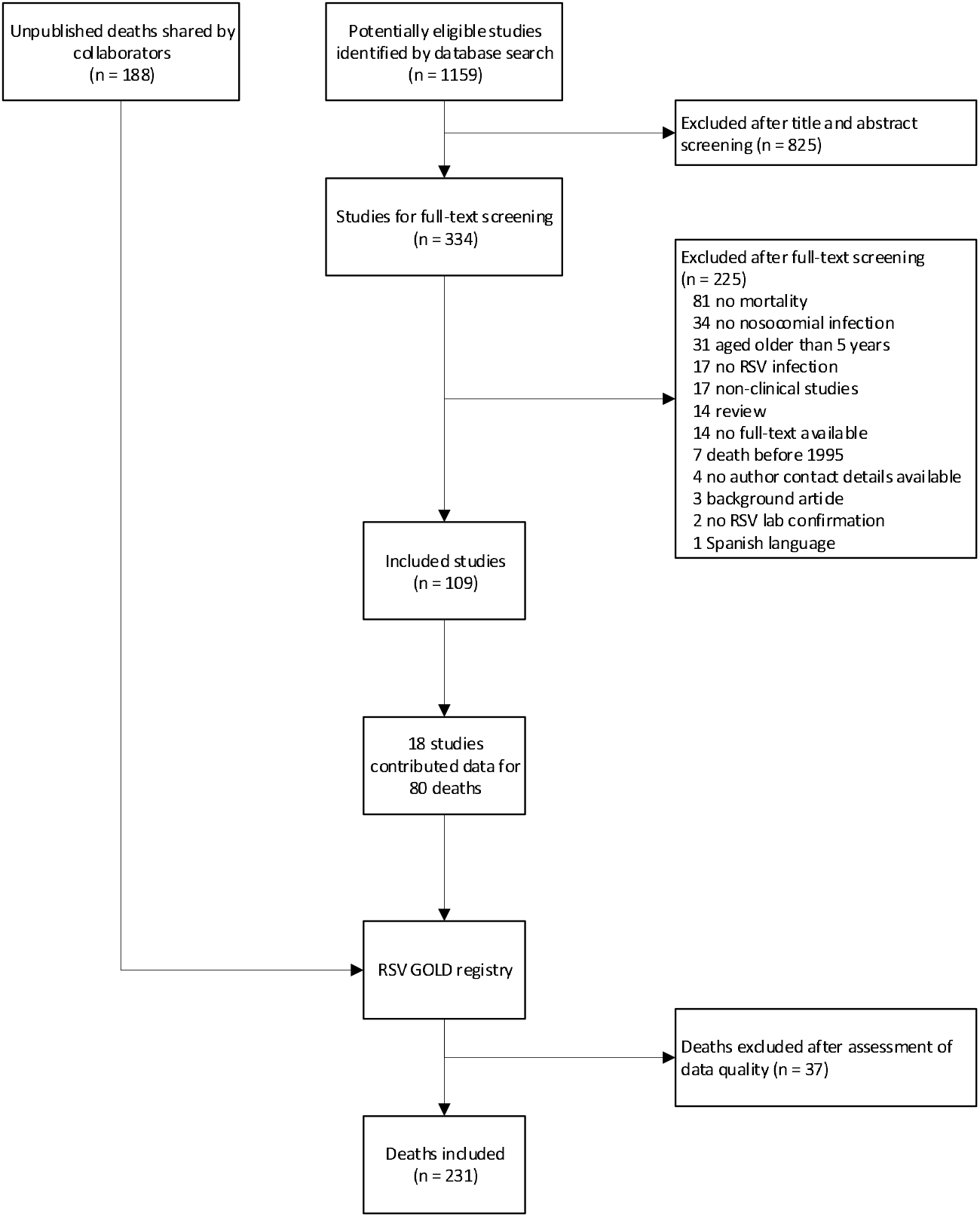
Flowchart of included nosocomial RSV deaths Abbreviations: RSV, respiratory syncytial virus.

**Figure 2:**
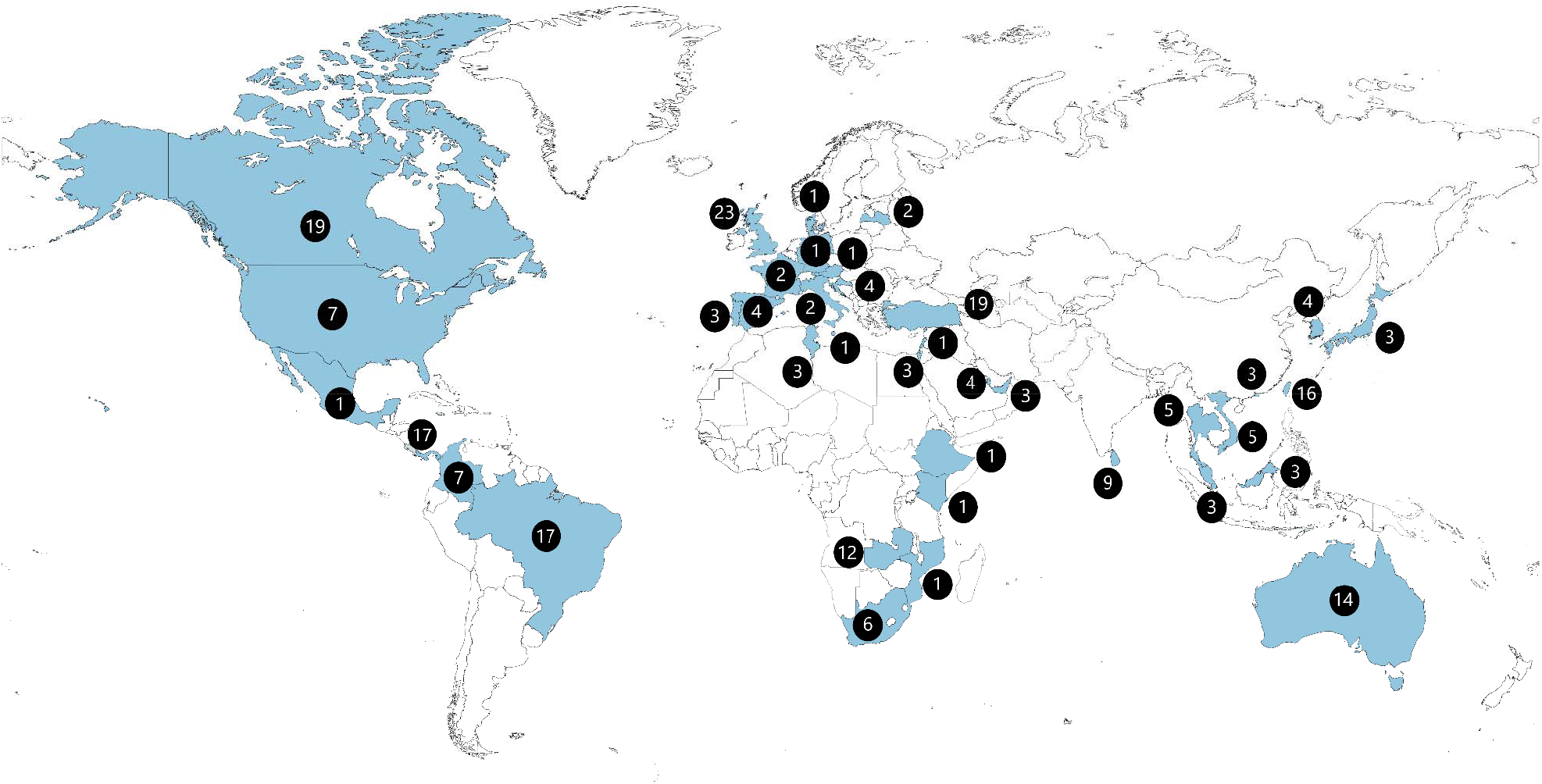
Location of death for children younger than 5 years with nosocomial RSV-related mortality included in the analysis Number of included deaths are given for each country (in blue) from which collaborators shared data.

### Clinical and demographic characteristics of children with nosocomial RSV-related mortality

The vast majority of children with nosocomial RSV-related death had one or more comorbidities (n=202, 87.4%) as expected. Congenital heart disease was the most prevalent comorbidity occurring in almost half (n=96, 41.6%) of all nosocomial deaths. In total, 106 (45.9%) children had been born prematurely. Median age at time of death was 5.4 (IQR: 2.5-12.1) months. Half of the deaths (51.9%) occurred within the first 6 months of life (Table 1, Figure 3).

**Table 1:** Characteristics of children younger than 5 years with nosocomial and community-acquired RSV-related in-hospital death

|  | **Nosocomial (n=231)** | **Community-acquired (n=931)** | **p value** |
| --- | --- | --- | --- |
| Male sex | 107 (46.3) | 500 (53.7) | 0.05 |
| Age at death (months) | 5.4 (2.5-12.1) | 6.0 (2.6-13.0) | 0.24 |
| Neonatal death | 18 (7.8) | 38 (4.1) | 0.03 |
| <3 months at death | 66 (28.6) | 244 (26.2) | 0.51 |
| <6 months at death | 120 (51.9) | 452 (48.5) | 0.38 |
| Year of death | 2012 (2006-2016) | 2011 (2006-2015) | 0.12 |
| LMIC | 32 (13.9) | 236 (25.3) | <0.001 |
| UMIC | 76 (32.9) | 338 (36.3) | 0.36 |
| HIC | 123 (53.2) | 357 (38.3) | <0.001 |
| Comorbidity§ | 202 (87.4) | 527 (56.6) | <0.001 |
| Congenital heart disease | 96 (41.6) | 209 (22.4) | <0.001 |
| Chronic lung disease | 42 (18.2) | 112 (12.0) | 0.02 |
| Genetic disease | 48 (20.8) | 134 (14.4) | 0.02 |
| Down syndrome | 18 (7.8) | 47 (5.0) | 0.11 |
| Neurological disease | 39 (16.9) | 135 (14.5) | 0.36 |
| Immune disorder | 15 (6.5) | 31 (3.3) | 0.04 |
| Other | 68 (29.4) | 139 (14.9) | <0.001 |
| Prematurity§ | 106 (45.9) | 225 (24.2) | <0.001 |
| Gestational age (weeks) | 36.0 (32.0-38.3); n=174 | 38.0 (34.0-39.0); n=496 | 0.004 |
| Birth weight (kgs) | 2.5 (1.7-3.1); n=168 | 2.7 (2.0-3.2); n=449 | 0.02 |
| Breastfeeding until age $\leq 4$ months | 36/139 (25.9) | 161/404 (39.9) | 0.003 |
| Length of hospital stay (days) | 23.0 (9.8-53.3); n=230 | 9.0 (4.0-20.0); n=901 | <0.001 |
| Intensive care unit (ICU) admission | 209/216 (96.8) | 641/868 (73.8) | <0.001 |
| Length of ICU stay (days) | 14.0 (7.0-30.0); n=194 | 10.0 (4.0-21.0); n=601 | <0.001 |
| Respiratory support | 210/210 (100) | 685/686 (99.9) | 1.00 |
| Mechanical ventilation | 186/230 (80.9) | 611/902 (67.7) | <0.001 |
| Duration of respiratory support (days) | 14.0 (7.0-29.0); n=195 | 10.0 (4.0-20.0); n=588 | <0.001 |

Data are presented as n (%), n/N (%), or median (IQR). §Considered absent when missing. Abbreviations: LMIC, low-income and lower-middle-income country; UMIC, upper middle-income country; HIC, high-income country.

**Figure 3:**
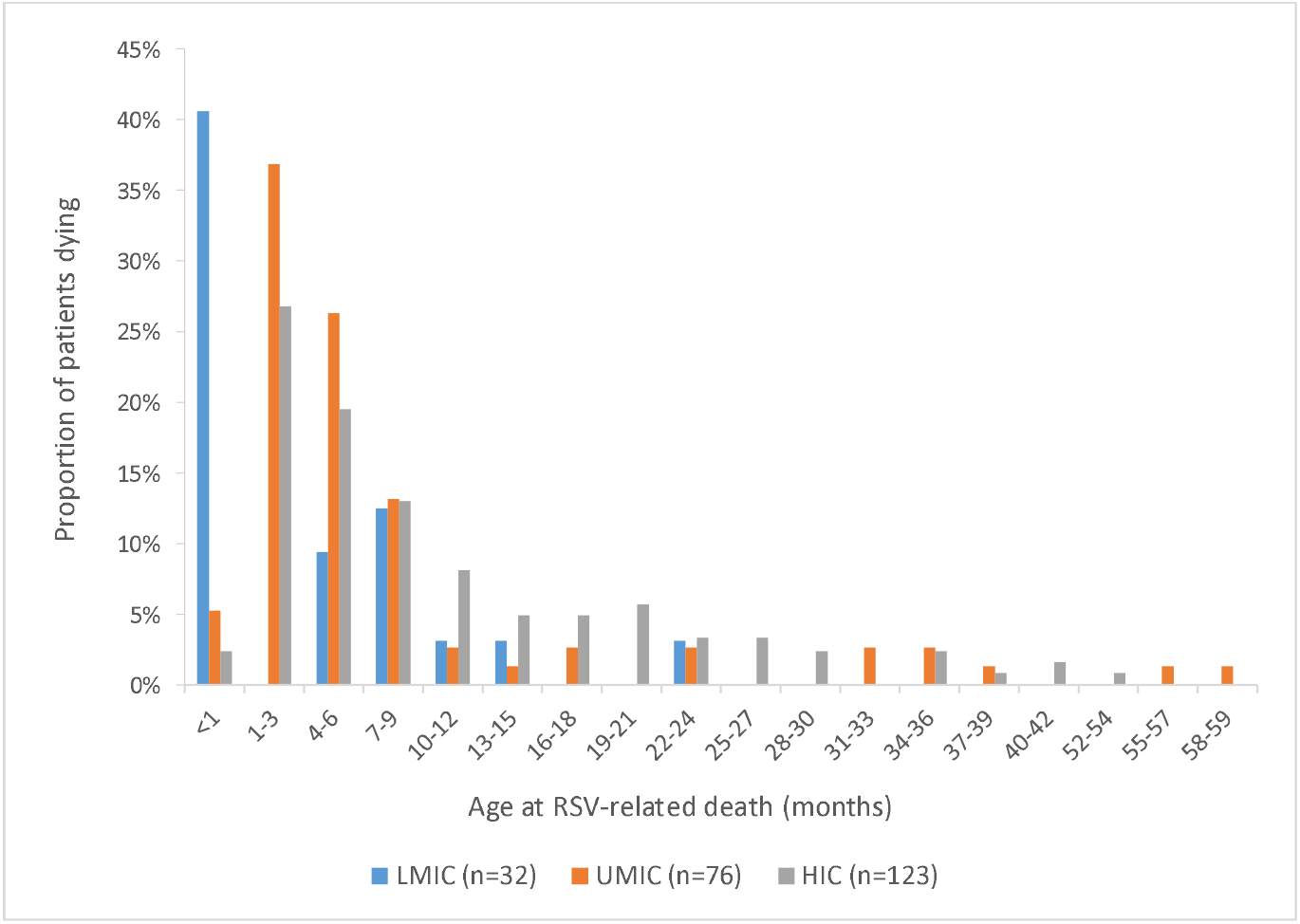
Age distribution at time of RSV-related nosocomial in-hospital death for children younger than 5 years from low- and middle-income countries (LMIC), upper-middle income countries (UMIC), and high-income countries (HIC)

### Nosocomial RSV-related mortality analyzed by World Bank Income group

Children from LMICs were younger at time of death than children from UMICs (1.5 vs. 5 months, p=0.003) and HICs (1.5 vs. 7 months, p<0.001). More children from LMICs died during the neonatal period (40.6%) than children from UMICs (3.9%) and HICs (1.6%), p<0.001. Furthermore, compared to HICs, children from LMICs were more often born prematurely (68.8% vs. 43.1%, p=0.01), had a shorter hospital stay (7 days vs. 37 days, p<0.001) and were less often mechanically ventilated (46.9% vs. 85.2%, p<0.001). Additional characteristics for children analyzed by income group are included in Supplemental Table 4.

### Nosocomial versus community-acquired RSV-related mortality

The proportion of preterm children was higher among children with nosocomial RSV infection compared to children with community-acquired RSV infection (45.9% vs. 24.2%, p<0.001). Likewise, a higher proportion of children with nosocomial RSV infection had at least one comorbidity (87.4% vs. 56.6%, p<0.001). Age at death was similar for both groups. We compared both nosocomial and community-acquired deaths for each income group separately to rule out sampling bias by underrepresentation of deaths from LMICs. In this analysis, children from LMICs with nosocomial RSV-related death were younger than children with community-acquired death (1.5 vs 5.0 months, p=0.002). For children from UMICs and HICs, age at death was similar (Supplemental Table 5-7). The odds ratios and 95% CIs for the presence of demographic and clinical characteristics in children with nosocomial vs. community-acquired RSV-related mortality in all included children and stratified by income group are presented in Supplemental Figure 1.

### Sensitivity analyses

When analyzing the data after excluding children with missing data for comorbidity or prematurity (n=19 for nosocomial RSV deaths; n=231 for community-acquired RSV deaths), our findings did not change (Supplemental Table 8). After excluding deaths collected from the post-mortem study in Zambia (n=12), median age increased to 5 months for nosocomial deaths from LMICs.

## DISCUSSION

Globally, RSV is a major cause of child mortality. Current RSV mortality burden estimates are largely based on community-acquired RSV-infections. We report the first global case series characterizing children younger than 5 years who died with nosocomial RSV infection. We observed that nosocomial RSV-related deaths accounted for 20% of all in-hospital RSV deaths reported to the RSV GOLD registry. This proportion indicates that nosocomial RSV-related deaths substantially contribute to the global burden of RSV-related child mortality.

While literature indicates that nosocomial infections generally occur at higher rates in LMICs than in HICs[3], the small number of nosocomial deaths from LMICs suggests these cases are underreported in LMICs. For example, data from the Zambia Pertussis Infant Mortality Estimation Study (ZPRIME) showed that 32% of RSV deaths in a tertiary referral hospital in Lusaka, Zambia, were due to nosocomial RSV infection.[18] The low number of LMIC nosocomial deaths in the RSV GOLD registry may be explained by sub-optimal testing capacity and a lack of active RSV surveillance systems and general underreporting of nosocomial deaths. Furthermore, ascertainment bias could play a role as data from LMICs mainly result from RSV studies that usually do not include nosocomial RSV deaths whereas data from HICs may also result from routine testing.

We found that age at death was similar for children with nosocomial and community-acquired RSV-related death. To our knowledge, there are no other studies available reporting on age in deceased pediatric patients with nosocomial and community-acquired infection. Two prospective studies from Vietnam and Germany report a younger age in pediatric patients with nosocomial RSV infection[8, 19], while studies from Brazil and the UK report an older age for nosocomial RSV patients (although the age difference was no longer significant when comparing comorbidity-matched controls in the UK study).[12, 20] More than half of reported nosocomial deaths were in children younger than 6 months at time of death. This age distribution indicates that infant-targeted RSV immunization strategies will prevent a large part of nosocomial RSV-related mortality globally.

Most children with nosocomial RSV-related mortality had, as expected, at least one comorbidity. Most frequently reported comorbidities were congenital heart disease, certain genetic disorders such as trisomy 21 (Down syndrome), and neurological disease. These findings are in line with findings from other studies that identified congenital heart disease, Down syndrome, immunodeficiency[6], and prematurity[8] as risk factors for acquiring RSV infection in the hospital. In a Canadian study, children with nosocomial RSV infection were more likely to have preexisting risk factors for severe disease including prematurity, immunodeficiency, lung disease, or heart disease, and stayed longer in the hospital than children with community-acquired RSV infection, which is comparable to our results.[9] In our registry, there were more premature infants with RSV-related nosocomial death from LMICs (69%) compared to UMICs (41%) or HICs (43%) and less children with comorbidity (69%) compared to children from HICs (96%). Possibly, in LMICs, children who are born preterm in the hospital may acquire nosocomial RSV infection and subsequently die at a younger age before comorbidities are identified. This contrasts with HICs, where children who die with RSV generally have comorbidities and are susceptible to severe nosocomial infection at an older age from repeated exposure to RSV through multiple hospital admissions. Taken together, differences in clinical characteristics may partially be explained by differences in quality of healthcare.

This study has several limitations inherent to retrospective data collection. First, the majority of the reported deaths were from UMICs and HICs while LMICs represent more than 75% of the global pediatric population and have a higher prevalence of nosocomial infections.[21] Second, since not all authors responded or shared data, we missed at least 24 deaths reported in the literature. All but three missed published deaths were from UMICs or HICs and median age at death was one month (information on age available for seven deaths). Third, authors employed variable definitions for nosocomial infection (time interval between admission and start of respiratory symptoms ranging from 48 hours to 7 days) which may have over- or underestimated our results. Although we did not collect these definitions, we checked for each hospital-acquired RSV mortality case whether the time interval between admission and symptoms was at least 48 hours. Furthermore, there were no studies that involved surveillance on nosocomial post-discharge deaths. The actual burden of nosocomial RSV-related mortality may therefore be higher than reported in the literature. Reporting bias could also lead to an underestimation of nosocomial RSV-related mortality as RSV outbreaks may not always be reported or detected by hospitals. Fourth, we did not collect information on the reason for hospital admission before the child acquired RSV, nor on location where RSV infection was acquired (ward or intensive care unit). Last, retrospective data vary in quality and completeness, although we attempted to limit the impact of this methodological weakness with data quality checks and direct verification with collaborators.

Our study has clinical implications. RSV is one of the major causes of nosocomial outbreaks in pediatric wards as it is highly transmissible, with a basic reproduction number - the number of secondary infections caused by one infected child - estimated between 1-9.[22] RSV is spread through direct contact or large droplet transmission but not through aerosol transmission.[23] Nosocomial RSV infections are associated with increased hospital costs due to prolonged hospitalization and extra interventions, and with high morbidity and mortality.[24, 25] Our results show that nosocomial RSV-related deaths may substantially contribute to the global burden of RSV-related child mortality. More than half of reported deaths were younger than 6 months and may have been prevented by infant-targeted RSV immunization strategies such as long-acting monoclonal antibodies. While the global community awaits market-approval and implementation of maternal vaccination and introduction of nirsevimab, a highly potent long-acting antibody that recently met the primary endpoint in a phase-III trial[11], infection-control measures remain of major importance to prevent nosocomial infections. Although high-quality evidence is lacking, multicomponent control strategies appear broadly successful.[26] Strict infection control measures including hand hygiene and the use of protective equipment (goggles, gloves, gowns, masks), can reduce nosocomial infection rates by 39-76%.[27]

In conclusion, this study provides insight into characteristics of global nosocomial RSV-related mortality in young children. Prospective studies and hospital-based surveillance are needed to demonstrate the magnitude of the nosocomial RSV burden in LMICs. Infection control measures, in particular hand hygiene, remain of significant importance to prevent nosocomial infections in pediatric wards worldwide.

## Data Availability

All data produced in the present study are available upon reasonable request to the authors.

## NOTES

### Funding

This work was supported by the Bill & Melinda Gates Foundation [OPP1148988].

### Potential conflicts of interest

LJB has regular interaction with pharmaceutical and other industrial partners. He has not received personal fees or other personal benefits. UMCU has received major funding (>€100,000 per industrial partner) for investigator-initiated studies from AbbVie, MedImmune, Janssen, the Bill and Melinda Gates Foundation, Nutricia (Danone) and MeMed Diagnostics. UMCU has received major cash or in kind funding as part of the public private partnership IMI-funded RESCEU project from GSK, Novavax, Janssen, AstraZeneca, Pfizer and Sanofi. UMCU has received major funding by Julius Clinical for participating in the INFORM study sponsored by MedImmune. UMCU has received minor funding for participation in trials by Regeneron and Janssen from 2015-2017 (total annual estimate less than €20,000). UMCU received minor funding for consultation and invited lectures by AbbVie, MedImmune, Ablynx, Bavaria Nordic, MabXience, Novavax, Pfizer, Janssen (total annual estimate less than €20,000). LJB is the founding chairman of the ReSViNET Foundation. HN has received grants from Innovative medicines initiative and Pfizer. He has received honorarium from Abbvie, Sanofi, ReViral, Janssen and Novavax.

## Acknowledgements

We are grateful to the collaborators of the RSV GOLD study group for sharing data with the RSV GOLD registry. We thank Dr. Sheikh Wasik Rahman, Mr. Shuborno Islam, Ms. Naito Kanon, Prof. Nawshad Ahmed, Dr. Rubana Sultana Aflatun and Dr. Sohel Mahmud of the Child Health Research Foundation for their intellectual, laboratory, and technical assistance with compiling the RSV dataset of Bangladesh. We thank Prachi Vora and Padmini Srikantiah for their scientific advice. Furthermore, we thank Femke Vernooij from the RSV GOLD student operations team for her help with data analyses.

## Notes

### Author Declarations

The institutional research board of the University Medical Centre Utrecht is the dedicated ethics oversight body for this study and waived the requirement for parental consent as only de-identified secondary patient data are used. Collaborators sharing data were encouraged to adhere to local site regulations for ethics approval. Where required, local ethical approval was obtained.

