## Supplementary File for "Nosocomial RSV-related in-hospital mortality in children <5 years: a global case series"

SUPPLEMENTARY APPENDIX

**Supplemental Table 1**: Literature search

| **Search terms** | **Date of search** |
| --- | --- |
| ("respiratory syncytial virus, human"[MeSH Terms] OR “RSV”[tiab] OR "Respiratory Syncytial Virus Infections"[Mesh] OR ("respiratory"[tiab] AND "syncytial"[tiab] AND "viruses"[tiab]) OR "respiratory syncytial viruses"[tiab] OR ("respiratory"[tiab] AND "syncytial"[tiab] AND "virus"[tiab]) OR "respiratory syncytial virus"[tiab] OR “bronchiolitis”[Mesh] or “bronchiolitis”[tiab]) AND ("Healthcare-Associated Pneumonia"[Mesh] OR “nosocomial”[tiab] OR “hospital-acquired” [Text Word] OR “nosocomial”[tiab] OR “in-hospital”[tiab] OR “hospital-borne”[tiab] OR “cross infection”[MeSH] OR “acquire*”[tiab]) | April 19, 2021 |

**Supplemental Table 2**: Publications from which data from nosocomial RSV deaths were contributed

| **PMID** | **Title** | **Authors** | **Journal** | **Year published** | **Reporting period** |
| --- | --- | --- | --- | --- | --- |
| 34845151 | Respiratory syncytial virus-attributable deaths in a major pediatric hospital in New South Wales, Australia, 1998-2018* | Saravanos GL et al. | Pediatr Infect Dis J | 2021 | January 1998 – December 2018 |
| 34472577 | Deaths attributed to respiratory syncytial virus in young children in high-mortality rate settings: report from Child Health and Mortality Prevention Surveillance (CHAMPS)* | Blau DM et al. | Clin Infect Dis | 2021 | December 2016 – December 2019 |
| 34472569 | Association of respiratory syncytial virus infection and underlying risk factors for death among young infants who died at University Teaching Hospital, Lusaka Zambia* | Forman LS et al. | Clin Infect Dis | 2021 | August 2017 – August 2020 |
| 32783380 | Risk factors of severe hospitalized respiratory syncytial virus infection in tertiary care center in Thailand | Aikphaibul P et al. | Influenza Other Respir Viruses | 2021 | January 2011 – December 2016 |
| 30552864 | First report of two consecutive respiratory syncytial virus outbreaks by the novel genotypes ON-1 and NA-2 in a neonatal intensive care unit | Silva DGBPD et al. | J Pediatr (Rio J) | 2020 | May 2013 – August 2013 |
| 32305431 | Clinical impact and direct costs of nosocomial  respiratory syncytial virus infections in the neonatal intensive care unit | Comas-García A et al. | Am J Infect Control | 2020 | July 2012 – December 2017 |
| 29788036 | Pediatric Investigators Collaborative Network on Infections in Canada Study of Respiratory Syncytial Virus-associated Deaths in Pediatric Patients in Canada, 2003-2013 | Tam J et al. | Clin Infect Dis | 2019 | 2003 - 2013 |
| 31521197 | Childhood nosocomial viral acute respiratory tract infections in teaching hospital Anuradhapura, Sri Lanka | Sampath Jayaweera JAA et al. | BMC Res Notes | 2019 | June 2013 – November 2014 |
| 25442868 | Risk factors associated with death in patients with severe respiratory syncytial virus infection | Lee YI et al. | J Microbio Immunol  Infect | 2016 | July 2001 – June 2010 |
| 25702707 | Characterization of hospital and community-acquired respiratory syncytial virus in children with severe lower respiratory tract infections in Ho Chi Minh City, Vietnam, 2010 | Tuan TA et al. | Influenza Other Respir Viruses | 2015 | January 2010 – December 2010 |
| 25179332 | Nosocomial respiratory syncytial virus infections in the palivizumab-prophylaxis era with implications regarding high-risk infants | Ashkenazi-Hoffnung L et al. | Am J Infect Control | 2014 | January 2008 – December 2010 |
| 24607551 | Nosocomial transmission of respiratory syncytial virus in an outpatient cancer center | Chu HY et al. | Biol Blood Marrow Transplant | 2014 | 2007 – 2008 and 2012 |
| 23897635 | Palivizumab prophylaxis during nosocomial outbreaks of respiratory syncytial virus in a neonatal intensive care unit: predicting effectiveness with an artificial neural network model | Saadah LM et al. | Pharmaco-therapy | 2014 | March – May 2005; December 2005 – January 2006; January – March 2006; April – July 2007 |
| 24564922 | Evaluation of respiratory syncytial virus group A and B genotypes among nosocomial and community-acquired pediatric infections in Southern Brazil | de-Paris F et al. | Virol J | 2014 | 2010 |
| 22415435 | Mortality and morbidity of nosocomial respiratory syncytial virus (RSV) infection in ventilated children--a ten year perspective | Thorburn K et al. | Minerva Anestesiol | 2012 | June 2001 – May 2011 |
| 21666538 | Hospital-acquired viral infection increases mortality in children with severe viral respiratory infection | Spaeder MC et al. | Pediatr Crit Care Med | 2011 | October 2002 – September 2008 |
| 20305582 | Nosocomial transmission of respiratory syncytial virus in neonatal intensive care and intermediate care units | Berger A et al. | Pediatr Infect Dis J | 2010 | January 2008 – March 2008 |
| 18653625 | Pre-existing disease is associated with a significantly higher risk of death in severe respiratory syncytial virus infection | Thorburn K | Arch Dis Child | 2009 | June 1999 – May 2007 |
| 18297695 | Molecular epidemiological analysis of a nosocomial outbreak of respiratory syncytial virus associated pneumonia in a kangaroo mother care unit in South Africa | Visser A et al. | J Med Virol | 2008 | March 2006 – May 2006 |
| 15236847 | RSV outbreak in a paediatric intensive care unit | Thorburn K et al. | J Hosp Infect | 2004 | October 2001 – March 2002 |
| *Studies were published after the systematic search; data had been shared before publication and these deaths are therefore displayed as unpublished deaths in Figure 1. 28 (35%) of included published deaths were due to an RSV outbreak. | | | | |  |

**Supplemental Table 3**: Origin of data for children younger than 5 years with RSV-related in-hospital death

| **Country** | **Nosocomial RSV-related death (N=231)** | **Community-acquired RSV-related death (N=931)** |
| --- | --- | --- |
| **Low-income** |  |  |
| Ethiopia | 1 (0.4%) | 0 |
| Burkina Faso | 0 | 2 (0.2%) |
| Mali | 0 | 12 (1.3%) |
| Mozambique | 1 (0.4%) | 6 (0.6%) |
| Uganda | 0 | 1 (0.1%) |
| **Lower-middle-income** |  |  |
| Bangladesh | 0 | 25 (2.7%) |
| Bolivia | 0 | 3 (0.3%) |
| India | 0 | 23 (2.5%) |
| Kenya | 1 (0.4%) | 59 (6.3%) |
| Morocco | 0 | 8 (0.9%) |
| Nicaragua | 0 | 17 (1.8%) |
| Pakistan | 0 | 18 (1.9%) |
| Philippines | 0 | 21 (2.3%) |
| Sri Lanka | 9 (3.9%) | 4 (0.4%) |
| Tunisia | 3 (1.3%) | 15 (1.6%) |
| Vietnam | 5 (2.2%) | 6 (0.6%) |
| Zambia | 12 (5.2%) | 16 (1.7%) |
| **Upper middle-income** |  |  |
| Argentina | 0 | 104 (11.2%) |
| Botswana | 0 | 4 (0.4%) |
| Brazil | 17 (7.4%) | 23 (2.5%) |
| Colombia | 7 (3.0%) | 25 (2.7%) |
| Indonesia | 0 | 14 (1.5%) |
| Jordan | 0 | 11 (1.2%) |
| Lebanon | 1 (0.4%) | 4 (0.4%) |
| Malaysia | 3 (1.3%) | 3 (0.3%) |
| Mexico | 1 (0.4%) | 13 (1.4%) |
| Panama | 17 (7.4%) | 27 (2.9%) |
| South Africa | 6 (2.6%) | 53 (5.7%) |
| Thailand | 5 (2.2%) | 30 (3.2%) |
| Turkey | 19 (8.2%) | 25 (2.7%) |
| **High-income** |  |  |
| Andorra | 0 | 1 (0.1%) |
| Australia | 14 (6.1%) | 24 (2.6%) |
| Austria | 1 (0.4%) | 1 (0.1%) |
| Canada | 19 (8.2%) | 53 (5.7%) |
| Chile | 0 | 3 (0.3%) |
| China | 0 | 2 (0.2%) |
| Croatia | 4 (1.7%) | 5 (0.5%) |
| Denmark | 1 (0.4%) | 3 (0.3%) |
| Finland | 0 | 3 (0.3%) |
| France | 2 (0.9%) | 6 (0.6%) |
| Germany | 1 (0.4%) | 6 (0.6%) |
| Greece | 0 | 14 (1.5%) |
| Hong Kong SAR, China | 3 (1.3%) | 12 (1.3%) |
| Israel | 3 (1.3%) | 13 (1.4%) |
| Italy | 2 (0.9%) | 1 (0.1%) |
| Japan | 3 (1.3%) | 18 (1.9%) |
| Korea, Rep. | 4 (1.7%) | 12 (1.3%) |
| Kuwait | 0 | 6 (0.6%) |
| Latvia | 2 (0.9%) | 6 (0.6%) |
| Malta | 1 (0.4%) | 1 (0.1%) |
| Netherlands | 0 | 10 (1.1%) |
| New Zealand | 0 | 3 (0.3%) |
| Poland | 0 | 1 (0.1%) |
| Portugal | 3 (1.3%) | 5 (0.5%) |
| Qatar | 4 (1.7%) | 8 (0.9%) |
| Singapore | 3 (1.3%) | 7 (0.8%) |
| Slovenia | 0 | 1 (0.1%) |
| Spain | 4 (1.7%) | 9 (1%) |
| Sweden | 0 | 5 (0.5%) |
| Switzerland | 0 | 1 (0.1%) |
| Taiwan | 16 (6.9%) | 22 (2.4%) |
| United Arab Emirates | 3 (1.3%) | 0 |
| United Kingdom | 23 (10%) | 33 (3.5%) |
| United States of America | 7 (3%) | 62 (6.7%) |
| Uruguay | 0 | 2 (0.2%) |

Abbreviations: RSV, respiratory syncytial virus.

**Supplemental Table 4**: Characteristics of children younger than 5 years with nosocomial RSV-related death categorized by income group

|  | **LMICs (n=32)** | **p value*** | **UMICs (n=76)** | **p value†** | **HICs (n=123)** | **p value‡** |
| --- | --- | --- | --- | --- | --- | --- |
| Male sex | 15 (46.9) | 0.83 | 39 (51.3) | 0.31 | 53 (43.1) | 0.84 |
| Age at death (months) | 1.5 (0.4-7.9); n=32 | 0.003 | 5.0 (2.1-8.0); n=76 | 0.004 | 7.0 (3.7-16.0); n=123 | <0.001 |
| Neonatal death | 13 (40.6) | <0.001 | 3 (3.9) | 0.37 | 2 (1.6) | <0.001 |
| <3 months at death | 19 (59.4) | 0.01 | 24 (31.6) | 0.04 | 23 (18.7) | <0.001 |
| <6 months at death | 23 (71.9) | 0.20 | 44 (57.9) | 0.06 | 53 (43.1) | 0.005 |
| Year of death | 2014 (2013-2019); n=32 | 0.17 | 2015 (2011-2018); n=76 | <0.001 | 2008 (2003-2013); n=123 | <0.001 |
| Comorbidity§ | 22 (68.8) | 0.20 | 62 (81.6) | 0.002 | 118 (95.9) | <0.001 |
| Congenital heart disease | 11 (34.4) | 0.50 | 21 (27.6) | 0.001 | 64 (52.0) | 0.11 |
| Chronic lung disease | 4 (12.5) | 0.48 | 6 (7.9) | 0.001 | 32 (26.0) | 0.16 |
| Genetic/chromosomal disease | 7 (21.9) | 0.59 | 13 (17.1) | 0.37 | 28 (22.8) | 1.00 |
| Down syndrome | 4 (12.5) | 0.48 | 6 (7.9) | 0.78 | 8 (6.5) | 0.27 |
| Neurological disease | 2 (6.3) | 0.22 | 13 (17.1) | 0.71 | 24 (19.5) | 0.11 |
| Immune disorder | 0 | 0.10 | 8 (10.5) | 0.27 | 7 (5.7) | 0.35 |
| Airway abnormality | 3 (9.4) | 0.36 | 3 (3.9) | 0.26 | 11 (8.9) | 1.00 |
| Malignancy | 0 | 0.10 | 7 (9.2) | 0.38 | 17 (13.8) | 0.02 |
| Liver disease | 0 | 0.32 | 5 (6.6) | 0.03 | 1 (0.8) | 1.00 |
| Renal disease | 0 | 0.32 | 5 (6.6) | 0.26 | 3 (2.4) | 1.00 |
| Pulmonary hypertension | 0 | N/A | 0 | 0.53 | 2 (1.6) | 1.00 |
| Biliary disease | 0 | 0.55 | 3 (3.9) | 0.16 | 1 (0.8) | 1.00 |
| Congenital abnormality | 1 (3.1) | 0.51 | 1 (1.3) | 1.00 | 1 (0.8) | 0.37 |
| Metabolic disorder | 0 | N/A | 0 | N/A | 0 | N/A |
| Endocrine disorder | 0 | 1.00 | 2 (2.6) | 0.64 | 2 (1.6) | 1.00 |
| Developmental disorder | 0 | N/A | 0 | 1.00 | 1 (0.8) | 1.00 |
| Other heart disease | 0 | N/A | 0 | N/A | 0 | N/A |
| Severe malnutrition | 0 | N/A | 0 | N/A | 0 | N/A |
| Gastro-intestinal disease | 0 | N/A | 0 | 0.16 | 5 (4.1) | 0.58 |
| Inflammatory disease | 0 | N/A | 0 | 1.00 | 1 (0.8) | 1.00 |
| Malaria | 0 | N/A | 0 | N/A | 0 | N/A |
| HIV/AIDS | 2 (6.3) | 0.09 | 0 | N/A | 0 | 0.04 |
| Tuberculosis | 0 | N/A | 0 | N/A | 0 | N/A |
| Prematurity§ | 22 (68.8) | 0.01 | 31 (40.8) | 0.77 | 53 (43.1) | 0.01 |
| Gestational age (weeks) | 35.0 (34.0-37.3); n=18 | 0.49 | 37.0 (32.0-39.0); n=61 | 0.63 | 36.0 (31.0-39.0); n=95 | 0.59 |
| Mother immunised during pregnancy | 11/16 (68.8) | 0.03 | 0/4 | 1.00 | 1/25 (4.0) | <0.001 |
| Birthweight (kgs) | 2.5 (2.0-2.8); n=20 | 0.48 | 2.7 (1.8-3.2); n=67 | 0.20 | 2.4 (1.3-3.1); n=81 | 0.74 |
| Exclusive breast feeding ≤4 months | 18/26 (69.2) | 0.001 | 14/53 (26.4) | 0.005 | 4/60 (6.7) | <0.001 |
| Length of hospital stay (days) | 6.5 (3.0-17.5); n=32 | <0.001 | 17.0 (10.0-41.0); n=75 | 0.004 | 37.0 (16.0-77.0); n=123 | <0.001 |
| Intensive care unit (ICU) admission | 16/19 (84.2) | 0.05 | 74/76 (97.4) | 0.64 | 119/121 (98.3) | 0.02 |
| Length of ICU stay (days) | 4.0 (2.0-17.0); n=15 | 0.02 | 12.0 (7.3-21.0); n=72 | 0.007 | 18.0 (8.0-40.0); n=107 | 0.002 |
| Respiratory support | 18/18 (100) | N/A | 74/74 (100) | N/A | 118/118 (100) | N/A |
| Mechanical ventilation | 15/32 (46.9) | <0.001 | 67/76 (88.2) | 0.67 | 104/122 (85.2) | <0.001 |
| Duration of respiratory support (days) | 3.0 (2.0-14.5); n=17 | 0.005 | 13.0 (8.0-18.8); n=72 | 0.03 | 18.0 (7.0-39.3); n=106 | <0.001 |
| Fever | 16/31 (51.6) | 0.83 | 30/62 (48.4) | 0.61 | 33/76 (43.4) | 0.52 |
| ≥1 other micro-organisms in respiratory sample | 10/29 (34.5) | 0.50 | 31/71 (43.7) | 0.45 | 55/111 (49.5) | 0.21 |
| ≥1 other micro-organisms in other samples | 6/20 (30) | 0.79 | 23/66 (34.8) | 0.007 | 46/79 (58.2) | 0.04 |
| Other children <12 years present in household | 8/14 (57.1) | 0.32 | 34/46 (73.9) | 0.13 | 48/80 (60.0) | 1.00 |
| Day care attendance | 1/10 (10) | 0.26 | 14/47 (29.8) | <0.001 | 4/72 (5.6) | 0.49 |
| Mother uneducated | 0/21 | 0.003 | 10/31 (32.3) | 0.006 | 1/31 (3.2) | 1.00 |
| Father uneducated | 0/17 | 0.12 | 4/23 (17.4) | 0.35 | 1/20 (5.0) | 1.00 |

Data are presented as n (%), n/N (%) or median (IQR). Statistical comparisons with χ^2^ exact test or Mann-Whitney U test with p values of less than 0.0167 taken to be significant according to the Bonferroni correction for multiple testing. Abbreviations: LMIC, low-income and lower-middle-income country; UMIC, upper middle-income country; HIC, high-income country; N/A, not available. *Low-income or lower-middle-income versus upper middle-income country. †Upper middle-income country versus high-income country. ‡Low-income or lower-middle-income country versus high-income country. §Considered absent when missing.

**Supplemental Figure 1.** Odds ratios and 95% confidence intervals for the presence of characteristics in children with nosocomial and community-acquired RSV-related in-hospital death in all included children and stratified by income group

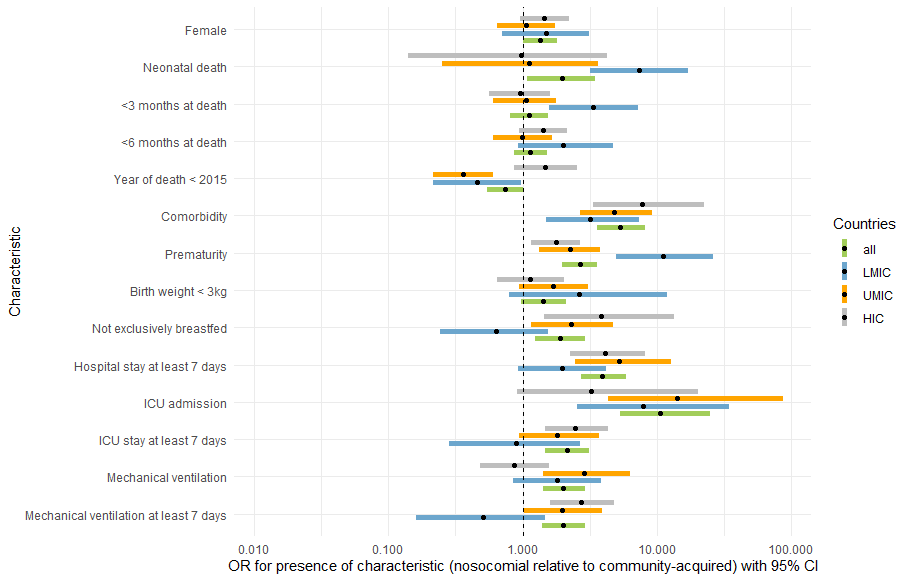

Odds ratio <1: characteristic more prevalent in children with community-acquired RSV-related in-hospital death. Odds ratio >1: characteristic more prevalent in children with nosocomial RSV-related in-hospital death.

**Supplemental Table 5**. Characteristics of children younger than 5 years with nosocomial and community-acquired RSV-related in-hospital death from LMICs

|  | **Nosocomial (n=32)** | **Community-acquired (n=236)** | **p value** |
| --- | --- | --- | --- |
| Male sex | 15 (46.9) | 134 (56.8) | 0.34 |
| Age at death (months) | 1.5 (0.4-7.9); n=32 | 5.0 (2.0-11.0); n=236 | 0.002 |
| Neonatal death | 13 (40.6) | 20 (8.5) | <0.001 |
| <3 months at death | 19 (59.4) | 72 (30.5) | 0.002 |
| <6 months at death | 23 (71.9) | 132 (55.9) | 0.13 |
| Year of death | 2014 (2013-2019); n=32 | 2012 (2009-2016); n=236 | 0.001 |
| Comorbidity§ | 22 (68.8) | 96 (40.7) | 0.004 |
| Congenital heart disease | 11 (34.4) | 36 (15.3) | 0.01 |
| Chronic lung disease | 4 (12.5) | 15 (6.4) | 0.26 |
| Genetic disease | 7 (21.9) | 8 (3.4) | 0.001 |
| Down syndrome | 4 (12.5) | 4 (1.7) | 0.008 |
| Neurological disease | 2 (6.3) | 13 (5.5) | 0.70 |
| Immune disorder | 0 | 1 (0.4) | 1.00 |
| Other | 6 (18.8) | 29 (12.3) | 0.40 |
| Prematurity§ | 22 (68.8) | 39 (16.5) | <0.001 |
| Gestational age (weeks) | 35.0 (34.0-37.3); n=18 | 38.0 (33.0-38.0); n=77 | 0.16 |
| Birth weight (kgs) | 2.5 (2.0-2.8); n=20 | 2.5 (1.8-3.1); n=82 | 0.75 |
| Breastfeeding until 4 months | 18/26 (69.2) | 67/114 (58.8) | 0.38 |
| Length of hospital stay (days) | 6.0 (3.0-17.5); n=32 | 4.0 (2.0-9.0); n=229 | 0.009 |
| Intensive care unit (ICU) admission | 16/19 (84.2) | 83/206 (40.3) | <0.001 |
| Length of ICU stay (days) | 4.0 (2.0-17.0); n=15 | 6.0 (2.0-11.0); n=94 | 0.94 |
| Respiratory support | 18/18 (100) | 100/101 (99.0) | 1.00 |
| Mechanical ventilation | 15 (46.9) | 76/233 (32.6) | 0.12 |
| Duration of respiratory support (days) | 3.0 (2.0-14.5); n=17 | 7.0 (2.0-13.0); n=83 | 0.65 |

Data are presented as n (%), n/N (%), or median (IQR). §Considered absent when missing. Abbreviations: LMIC, low-income and lower-middle-income country.

**Supplemental Table 6**. Characteristics of children younger than 5 years with nosocomial and community-acquired RSV-related in-hospital death from UMICs

|  | **Nosocomial (n=76)** | **Community-acquired (n=338)** | **p value** |
| --- | --- | --- | --- |
| Male sex | 39 (51.3) | 179 (53.0) | 0.80 |
| Age at death (months) | 5.0 (2.1-8.0); n=76 | 4.7 (2.0-9.0); n=338 | 0.81 |
| Neonatal death | 3 (3.9) | 12 (3.6) | 0.74 |
| <3 months at death | 24 (31.6) | 103 (30.5) | 0.89 |
| <6 months at death | 44 (57.9) | 196 (58.0) | 1.00 |
| Year of death | 2015 (2011-2018); n=76 | 2011 (2005-2016); n=338 | <0.001 |
| Comorbidity§ | 62 (81.6) | 162 (47.9) | <0.001 |
| Congenital heart disease | 21 (27.6) | 63 (18.6) | 0.08 |
| Chronic lung disease | 6 (7.9) | 45 (13.3) | 0.25 |
| Genetic disease | 13 (17.1) | 33 (9.8) | 0.07 |
| Down syndrome | 6 (7.9) | 22 (6.5) | 0.62 |
| Neurological disease | 13 (17.1) | 35 (10.4) | 0.11 |
| Immune disorder | 8 (10.5) | 10 (3.0) | 0.008 |
| Other | 21 (27.6) | 38 (11.2) | 0.001 |
| Prematurity§ | 31 (40.8) | 79 (23.4) | 0.004 |
| Gestational age (weeks) | 37.0 (32.0-39.0); n=61 | 38.0 (35.0-39.0); n=169 | 0.13 |
| Birth weight (kgs) | 2.7(1.8-3.2); n=67 | 2.8 (2.3-3.2); n=164 | 0.08 |
| Breastfeeding until 4 months | 14/53 (26.4) | 60/133 (45.1) | 0.02 |
| Length of hospital stay (days) | 17.0 (10.0-41.0); n=75 | 10.0 (5.0-19.8); n=332 | <0.001 |
| Intensive care unit (ICU) admission | 74/76 (97.4) | 225/311 (72.3) | <0.001 |
| Length of ICU stay (days) | 12.0 (7.3-21.0); n=72 | 11.0 (6.0-22.0); n=196 | 0.51 |
| Respiratory support | 74/74 (100) | 249/249 (100) | N/A |
| Mechanical ventilation | 67/76 (88.2) | 232/321 (72.3) | 0.003 |
| Duration of respiratory support (days) | 13.0 (8.0-18.8); n=72 | 10.0 (6.0-21.0); n=216 | 0.25 |

Data are presented as n (%), n/N (%), or median (IQR). §Considered absent when missing. Abbreviations: UMIC, upper middle-income country; N/A, not available.

**Supplemental Table 7**. Characteristics of children younger than 5 years with nosocomial and community-acquired RSV-related in-hospital death from HICs

|  | **Nosocomial (n=123)** | **Community-acquired (n=357)** | **p value** |
| --- | --- | --- | --- |
| Male sex | 53 (43.1) | 187 (52.4) | 0.09 |
| Age at death (months) | 7.0 (3.7-16.0); n=123 | 9.9 (3.9-22.0); n=357 | 0.06 |
| Neonatal death | 2 (1.6) | 6 (1.7) | 1.00 |
| <3 months at death | 23 (18.7) | 69 (19.3) | 1.00 |
| <6 months at death | 53 (43.1) | 124 (34.7) | 0.11 |
| Year of death | 2008 (2003-2013); n=123 | 2010 (2005-2014); n=357 | 0.05 |
| Comorbidity§ | 118 (95.9) | 269 (75.4) | <0.001 |
| Congenital heart disease | 64 (52.0) | 110 (30.8) | <0.001 |
| Chronic lung disease | 32 (26.0) | 52 (14.6) | 0.006 |
| Genetic disease | 28 (22.8) | 93 (26.1) | 0.55 |
| Down syndrome | 8 (6.5) | 21 (5.9) | 0.83 |
| Neurological disease | 24 (19.5) | 87 (24.4) | 0.32 |
| Immune disorder | 7 (5.7) | 20 (5.6) | 1.00 |
| Other | 41 (33.3) | 72 (20.2) | 0.004 |
| Prematurity§ | 53 (43.1) | 107 (30.0) | 0.01 |
| Gestational age (weeks) | 36.0 (31.0-39.0); n=95 | 37.0 (34.0-39.0); n=250 | 0.05 |
| Birth weight (kgs) | 2.4 (1.3-3.1); n=81 | 2.6 (1.8-3.2); n=203 | 0.13 |
| Breastfeeding until 4 months | 4/60 (6.7) | 34/157 (21.7) | 0.009 |
| Length of hospital stay (days) | 37.0 (16.0-77.0); n=123 | 12.0 (5.0-28.0); n=340 | <0.001 |
| Intensive care unit (ICU) admission | 119/121 (98.3) | 333/351 (94.9) | 0.12 |
| Length of ICU stay (days) | 18.0 (8.0-40.0); n=107 | 10.0 (4.0-23.0); n=311 | <0.001 |
| Respiratory support | 118/118 (100) | 336/336 (100) | N/A |
| Mechanical ventilation | 104/122 (85.2) | 303/348 (87.1) | 0.64 |
| Duration of respiratory support (days) | 18.0 (7.0-39.3); n=106 | 10.0 (4.0-22.0); n=289 | <0.001 |

Data are presented as n (%), n/N (%), or median (IQR). §Considered absent when missing. Abbreviations: HIC, high-income country; N/A, not available.

**Supplemental Table 8**. Characteristics of children younger than 5 years with nosocomial and community-acquired RSV-related in-hospital death, excluding children with missing data for comorbidity or prematurity

|  | **Nosocomial (n=212)** | **Community-acquired (n=700)** | **p value** |
| --- | --- | --- | --- |
| Male sex | 98 (46.2) | 382 (54.6) | 0.03 |
| Age at death (months) | 6.0 (2.9-12.0); n=212 | 6.0 (2.5-12.3); n=700 | 0.88 |
| Neonatal death | 12 (5.7) | 26 (3.7) | 0.24 |
| <3 months at death | 55 (25.9) | 194 (27.7) | 0.66 |
| <6 months at death | 107 (50.5) | 347 (49.6) | 0.88 |
| Year of death | 2012 (2007-2016); n=212 | 2011 (2006-2015); n=700 | 0.51 |
| LMIC | 24 (11.3) | 118 (16.9) | 0.05 |
| UMIC | 70 (33.0) | 275 (39.3) | 0.11 |
| HIC | 118 (55.7) | 307 (43.9) | 0.003 |
| Comorbidity | 194 (91.5) | 456 (65.1) | <0.001 |
| Congenital heart disease | 93/202 (46.0) | 191/633 (30.2) | <0.001 |
| Chronic lung disease | 40/200 (20.0) | 106/620 (17.1) | 0.34 |
| Genetic disease | 45/201 (22.4) | 118/602 (19.6) | 0.42 |
| Down syndrome | 17/173 (9.8) | 43/472 (9.1) | 0.76 |
| Neurological disease | 36/202 (17.8) | 114/612 (18.6) | 0.84 |
| Immune disorder | 14/199 (7.0) | 28/613 (4.6) | 0.20 |
| Other | 63 (29.7) | 114 (16.3) | <0.001 |
| Prematurity | 98 (46.2) | 209 (29.9) | <0.001 |
| Gestational age (weeks) | 36.0 (32.0-38.8); n=172 | 38.0 (34.0-39.0); n=483 | 0.005 |
| Birth weight (kgs) | 2.5 (1.7-3.1); n=162 | 2.7 (2.0-3.2); n=423 | 0.05 |
| Breastfeeding until 4 months | 31/130 (23.8) | 141/368 (38.3) | 0.003 |
| Length of hospital stay (days) | 23.0 (10.0-62.0); n=211 | 10.0 (5.0-22.0); n=680 | <0.001 |
| Intensive care unit (ICU) admission | 198/204 (97.1) | 566/683 (82.9) | <0.001 |
| Length of ICU stay (days) | 14.0 (8.0-31.0); n=184 | 10.0 (4.8-22.0); n=530 | <0.001 |
| Respiratory support | 200/200 (100) | 587/587 (100) | N/A |
| Mechanical ventilation | 177/211 (83.9) | 537/674 (79.7) | 0.20 |
| Duration of respiratory support (days) | 14.0 (7.0-30.0); n=186 | 10.0 (5.0-20.0); n=520 | <0.001 |

Data are presented as n (%), n/N (%), or median (IQR). Abbreviations: LMIC, low-income and lower-middle-income country; UMIC, upper middle-income country; HIC, high-income country; N/A, not available.

**Supplemental Figure 2:** Age distribution at time of RSV-related nosocomial in-hospital death for healthy term children, premature children, and term children with comorbidities

Proportion of patients dying

| Age at RSV-related death (months) | healthy term (n=6) | healthy preterm (n=23) | comorbidity (n=202) |
| --- | --- | --- | --- |
| <1 | 16.7% | 26.1% | 6.4% |
| 1-3 | 16.7% | 60.9% | 26.2% |
| 4-6 | 16.7% | 8.7% | 21.8% |
| 7-9 | 16.7% | 4.3% | 13.9% |
| 10-12 | 0% | 0% | 6.4% |
| 13-15 | 0% | 0% | 4% |
| 16-18 | 0% | 0% | 5% |
| 19-21 | 0% | 0% | 3.5% |
| 22-24 | 0% | 0% | 3.5% |
| 25-27 | 0% | 0% | 2% |
| 28-30 | 0% | 0% | 1.5% |
| 31-33 | 0% | 0% | 0.5% |
| 34-36 | 0% | 0% | 2.5% |
| 37-39 | 0% | 0% | 1% |
| 40-42 | 0% | 0% | 1% |
| 52-54 | 0% | 0% | 0.5% |
| 55-57 | 0% | 0% | 0% |
| 58-59 | 0% | 0% | 0.5% |
